# Effect of the COVID-19 pandemic and maternal SARS-CoV-2 status on breastfeeding practices in the COMBO cohort

**DOI:** 10.1101/2024.07.14.24309868

**Authors:** Lauren Walzer, Presley Nichols, Monica K. Amoo-Achampong, Melissa E. Glassman, Margaret H. Kyle, Maha Hussain, Wanda Abreu, Tessa Scripps, Adrita Khan, Erin Hanft, Ruiyang L. Xu, Mary Ann LoFrumento, Anglina Kataria, Melissa S. Stockwell, Dani Dumitriu, Cristina R. Fernández

## Abstract

**Objectives:** To investigate the role of prenatal maternal SARS-CoV-2 infection and delivery during the COVID-19 pandemic on breastfeeding practices through 6 months postpartum.

**Study design:** A cohort of mother-infant dyads who delivered at a large medical center in New York City and who had either documented history of prenatal SARS-CoV-2 infection, no history of SARS-CoV-2 infection, or who delivered in February 2020 prior to the COVID-19 pandemic were invited to participate in the COVID-19 Mother Baby Outcomes (COMBO) Initiative. These three groups of mothers completed surveys about their breastfeeding practices at 1, 2, 4, and/or 6 months postpartum, and responses were compared across enrollment group at each postpartum assessment period.

**Results:** Data from 557 mother-infant dyads was included. The SARS-CoV-2 infected group reported significantly lower rates of mostly or exclusively breastfeeding at 1 month, 2 months, and 4 months postpartum. When analyses were separated by birth timing and maternal ethnicity, these differences between infected and uninfected mothers were only significant in the second pandemic wave delivery group and in Latina mothers.

**Conclusions for practice:** Maternal SARS-CoV-2 infection during pregnancy was associated with lower rates of mostly or exclusively breastfeeding through 4 months postpartum. These differences may have been driven by delivery timing during the pandemic and self-reported maternal ethnicity. These results suggest a need to increase resources and support for breastfeeding during healthcare-disruptive events such as the pandemic, particularly for those belonging to groups historically underserved in healthcare.

## Introduction

Breastfeeding has numerous benefits for mothers and babies.^1^ Support for breastfeeding usually begins with hospital maternity and newborn care services. Breastfeeding rates in the US have risen in the last decade^2^ with increased support for maternity and newborn care championing breastfeeding.^3^ Peer and professional support, both in hospital and after discharge, may also promote breastfeeding.^4^

The COVID-19 pandemic disrupted hospital-based care models and traditional professional and peer breastfeeding support networks, with ramifications for breastfeeding success. Risk for mother-to-newborn transmission of SARS-CoV-2 was not well understood early in the pandemic, and initial breastfeeding guidance from health organizations was mixed.^5^ Several organizations initially recommended separating SARS-CoV-2 positive mothers from their babies, and many hospitals reduced or barred postpartum unit visitors, newborn rooming-in, and in-person lactation support. Changes to newborn outpatient care practices,^6^ social distancing, lockdowns and other stressors may have altered traditional provider, family, and peer networks of support.^7,8,9^

New York City (NYC) was at the epicenter of the US pandemic’s first wave from March to May 2020, with rates of polymerase chain reaction (PCR)-positivity in laboring women peaking at 15.3% in the first two weeks of universal testing at admission starting March 22, 2020.^10^ We previously reported on the first babies born to mothers with COVID-19 at our NYC-based hospital system, showing no clinical evidence of vertical transmission despite rooming-in and encouraging direct breastfeeding with appropriate infection control measures.^11^ Given the importance of hospital maternity and newborn care practice and support networks in breastfeeding initiation and success, this study sought to characterize the role of maternal SARS-CoV-2 infection and birth during the COVID-19 pandemic on breastfeeding outcomes through 6 months of age.

## Methods

### Population

This study is part of Columbia University’s COVID-19 Mother Baby Outcomes (COMBO) Initiative, an on-going longitudinal study established in May 2020 to monitor the health of mother-infant dyads with and without history of prenatal SARS-CoV-2 infection. Dyads who delivered at the Columbia University Irving Medical Center-affiliated NewYork-Presbyterian (NYP) Morgan Stanley Children’s Hospital and NewYork-Presbyterian Allen Hospital were enrolled. Enrollment strategies for COMBO have varied. During the period of this study, all mothers who delivered since the onset of the pandemic in NYC with electronic health record (EHR) documentation of SARS-CoV-2 infection (exposed group) during pregnancy were invited to participate. For each enrolled dyad in the exposed group, 1-3 case-matched dyads with no documented history of SARS-CoV-2 infection during pregnancy were enrolled based on infant gestational age, sex, delivery mode, and birth date within a 1-2 week window. An additional group of dyads delivering at these hospitals in February 2020, prior to the COVID-19 pandemic onset in NYC, were recruited to COMBO as a pre-pandemic birth-stress control group.

### Study design

We report on breastfeeding practices at 1, 2, 4, and 6 months postpartum of 557 dyads enrolled in COMBO with birthdates between February 1, 2020, and April 30, 2021. While COMBO’s overarching design is a prospective case-matched cohort study, dyads can enter the study at any time during pregnancy or postpartum, and therefore, data reported here is statistically treated as cross-sectional for each evaluated time-point due to few dyads (n = 71, 12.7%) contributing data for all time-points. Mothers were approached by telephone and e-mail and gave their informed consent prior to enrollment in COMBO as a mother-infant dyad. Mothers completed questionnaires (Table 1) about their breastfeeding practices in English or Spanish either over the phone or online secure survey in REDCap (version 12.0.29). All procedures with human subjects were approved by the institutional review board of Columbia University Irving Medical Center and have therefore been performed in accordance with the ethical standards laid down in the 1964 Declaration of Helsinki and its later amendments.

**Table 1.** Breastfeeding-related variables collected by maternal self-report.

| **Maternal self-reported**<br>**breastfeeding-related questions<sup>a</sup>** | **SARS-CoV-2 Negative**<br>**(N=267)** |  |  |  | **SARS-CoV-2 Positive**<br>**(N=221)** |  |  |  |
| --- | --- | --- | --- | --- | --- | --- | --- | --- |
|  | **1 month**<br>**(n=125)** | **2 months**<br>**(n=181)** | **4 months<sup>b</sup>**<br>**(n=189)** | **6 months<sup>c</sup>**<br>**(n=156)** | **1 month**<br>**(n=92)** | **2 months**<br>**(n=126)** | **4 months<sup>b</sup>**<br>**(n=122)** | **6 months<sup>c</sup>**<br>**(n=124)** |
| **How are you feeding the baby?** |  |  |  |  |  |  |  |  |
| All breastmilk | 55 (44%) | 34 (19%) | 14 (7%) | 46 (29%) | 29 (32%) | 17 (13%) | 12 (10%) | 31 (25%) |
| Mostly breastmilk but some formula | 27 (22%) | 12 (7%) | 8 (4%) | 22 (14%) | 18 (20%) | 11 (9%) | 8 (7%) | 17 (14%) |
| Equal amounts of breastmilk and formula | 10 (8%) | 6 (3%) | 3 (2%) | 10 (6%) | 10 (11%) | 7 (6%) | 3 (2%) | 4 (3%) |
| Mostly formula but some breastmilk | 21 (17%) | 10 (5%) | 5 (3%) | 10 (6%) | 23 (25%) | 9 (7%) | 5 (4%) | 10 (8%) |
| All formula | 12 (10%) | 10 (5%) | 16 (8%) | 68 (44%) | 12 (13%) | 15 (12%) | 13 (11%) | 62 (50%) |
| **Has it always been your plan to do mostly/all breastmilk?** |  |  |  |  |  |  |  |  |
| Yes | 74 (59%) | 44 (24%) | 22 (12%) | 66 (42%) | 42 (46%) | 25 (20%) | 20 (16%) | 46 (37%) |
| No | 4 (3%) | 1 (0.6%) | - | 2 (1%) | 4 (4%) | - | - | 1 (0.8%) |
| I don't know | 2 (2%) | 1 (0.6%) | - | 1 (0.6%) | 1 (1%) | - | - | 1 (0.8%) |
| **How would you rate your experience surrounding education on breastfeeding during your hospital stay?** |  |  |  |  |  |  |  |  |
| outstanding | 12 (10%) | 11 (6%) | 6 (3%) | - | 8 (9%) | 8 (6%) | 4 (3%) | - |
| good | 37 (30%) | 25 (14%) | 15 (8%) | - | 38 (41%) | 16 (13%) | 14 (11%) | - |
| neutral | 46 (37%) | 12 (7%) | 3 (2%) | - | 20 (22%) | 12 (10%) | 2 (2%) | - |
| Poor | 10 (8%) | 9 (5%) | 3 (2%) | - | 9 (10%) | 4 (3%) | 2 (2%) | - |
| Very poor | 7 (6%) | 3 (2%) | 3 (2%) | - | 5 (5%) | 1 (0.7%) | 6 (5%) | - |
| **How would you rate the hands-on support you received from staff regarding breastfeeding (i.e. helping the baby to latch, showing you how to hand express or pump)?** |  |  |  |  |  |  |  |  |
| outstanding | 23 (18%) | 15 (8%) | 7 (4%) | - | 16 (17%) | 11 (9%) | 3 (2%) | - |
| good | 39 (31%) | 24 (13%) | 15 (8%) | - | 33 (36%) | 14 (11%) | 14 (11%) | - |
| neutral | 33 (26%) | 9 (5%) | 4 (2%) | - | 17 (18%) | 9 (7%) | 3 (2%) | - |
| Poor | 12 (10%) | 7 (4%) | 2 (1%) | - | 7 (8%) | 2 (1%) | 3 (2%) | - |
| Very poor | 6 (5%) | 6 (3%) | 2 (1%) | - | 7 (8%) | 4 (3%) | 5 (4%) | - |
| **How would you rate communication from healthcare providers with respect to breastfeeding?** |  |  |  |  |  |  |  |  |
| outstanding | 20 (16%) | 13 (7%) | 6 (3%) | - | 13 (14%) | 9 (7%) | 5 (4%) | - |
| good | 45 (36%) | 23 (13%) | 13 (7%) | - | 36 (39%) | 19 (15%) | 12 (10%) | - |
| neutral | 36 (29%) | 16 (9%) | 6 (3%) | - | 20 (22%) | 6 (5%) | 4 (3%) | - |
| Poor | 8 (6%) | 5 (3%) | 3 (2%) | - | 7 (8%) | 5 (4%) | 4 (3%) | - |
| Very poor | 4 (3%) | 4 (2%) | 2 (1%) | - | 4 (4%) | 3 (2%) | 3 (2%) | - |
| **Were you seen by a Lactation Consultant prior to hospital discharge?** |  |  |  |  |  |  |  |  |
| Yes | 84 (67%) | 48 (27%) | 21 (11%) | - | 48 (52%) | 19 (15%) | 17 (14%) | - |
| No | 28 (22%) | 12 (7%) | 8 (4%) | - | 30 (33%) | 21 (17%) | 11 (9%) | - |
| I don't know | - | - | 1 (0.5%) | - | - | - | - | - |
| **When you left the hospital, did you feel confident about knowing how to feed your baby once home?** |  |  |  |  |  |  |  |  |
| Completely confident | 61 (49%) | 31 (17%) | 24 (13%) | - | 50 (54%) | 42 (33%) | 27 (22%) | - |
| Somewhat confident | 43 (34%) | 25 (14%) | 15 (8%) | - | 30 (33%) | 9 (7%) | 7 (6%) | - |
| A little bit confident | 15 (12%) | 11 (6%) | 3 (2%) | - | 9 (10%) | 3 (2%) | 4 (3%) | - |
| Not at all confident | 5 (4%) | 5 (3%) | 4 (2%) | - | 2 (2%) | 4 (3%) | 3 (2%) | - |
| Don't remember | 1 (0.8%) | - | - | - | - | - | - | - |
| **Have you had any of the following breastfeeding problems or concerns since arriving home from the hospital? (select all that apply)** |  |  |  |  |  |  |  |  |
| Concerns about milk supply | 49 (39%) | 26 (14%) | 11 (6%) | - | 29 (32%) | 14 (11%) | 12 (10%) | - |
| Difficulties with latch | 42 (34%) | 12 (7%) | 5 (3%) | - | 21 (23%) | 8 (6%) | 4 (3%) | - |
| Breast pain/engorgement | 40 (32%) | 19 (10%) | 12 (6%) | - | 19 (21%) | 14 (11%) | 5 (4%) | - |
| Nipple soreness/trauma | 50 (40%) | 20 (11%) | 13 (7%) | - | 26 (28%) | 10 (8%) | 8 (7%) | - |
| Demands of breastfeeding being too overwhelming | 30 (24%) | 13 (7%) | 8 (4%) | - | 18 (20%) | 3 (2%) | 8 (7%) | - |
| Other<sup>d</sup> | 11 (9%) | 4 (2%) | 4 (2%) | - | 5 (5%) | 4 (3%) | 2 (2%) | - |

<sup>a</sup>Due to missing data not all variables sum to 100%
<sup>b</sup>4 month totals excludes February Birth-Stress controls
<sup>c</sup>Infants enrolled into COMBO Study at 6 months of age only had questions about infant feeding type and maternal prenatal breastfeeding intention asked on their baseline intake questionnaire
<sup>d</sup>Breastfeeding problems or concerns written in by mothers who selected 'Other' include: concerns about not producing enough milk/baby not being full after breastfeeding, concerns about baby not wanting to latch, pain with letdown, mastitis and clogged milk ducts, oversupply, exhaustion/lack of energy, infant spit up/colic, newborn tongue tie, baby latching well onto one breast but not the other breast, concern about baby developing a food allergy against something mother is eating

### Maternal SARS-CoV-2 Infection Status

Our main predictor variable was maternal prenatal SARS-CoV-2 infection status. All delivering mothers at the NYP hospitals on or after March 22, 2020 were tested via nasopharyngeal PCR for SARS-CoV-2 at delivery. Beginning July 20, 2020, all mothers were also tested for antibodies to SARS-CoV-2. For the period prior to November 1, 2020 and mothers for whom conception was calculated to have occurred after the onset of the pandemic in NYC (defined as March 1, 2020, when the first patient was diagnosed), maternal prenatal SARS-CoV-2 infection status was determined by the presence of a positive SARS-CoV-2 nasopharyngeal PCR test or serology test in the maternal EHR. For the period after November 1, 2020, maternal EHRs were also assessed for all other SARS-CoV-2 testing during pregnancy that occurred within the NYP system. For each mother identified as having SARS-CoV-2 infection during pregnancy, the EHR was reviewed to determine the date of symptom onset, which was then referenced to the infant’s gestational age to determine the trimester of pregnancy in which infection occurred.

### Birth timing during the COVID-19 pandemic

Our secondary predictor variable was infant birth timing during the COVID-19 pandemic. Birth timing was categorized as: March 1 through August 31, 2020 (First wave of pandemic in NYC), and September 1, 2020 through April 30, 2021 (Second wave of pandemic in NYC).

### Maternal ethnicity

Our tertiary predictor variable was self-reported maternal ethnicity. Each mother self-identified in the EHR as “Hispanic or Latino or Spanish Origin”, herein defined as “Latina”, or as “Not Hispanic or Latino or Spanish Origin”, herein defined as “Not Latina”.

### Breastfeeding Practice

Our main outcome variable was breastfeeding practice, which was measured from responses to an over the phone or electronically self-administered survey. We used a question previously used by our group in published work^12^ and available in both English and Spanish: “How are you feeding the baby?”. Response categories included: *all breastmilk*, *mostly breastmilk but some formula*, *equal amounts breastmilk and formula, mostly formula but some breastmilk* and *all formula*. Secondary outcome variables were maternal responses to seven additional questions related to breastfeeding practices developed by our group to capture information particularly relevant during the pandemic. All breastfeeding-related questions are presented in Table 1.

### Covariates

Maternal and infant sociodemographic characteristics and in-hospital newborn feeding data were abstracted from the EHR.

### Statistical Analyses

Bivariate analyses compared maternal and newborn factors by maternal prenatal SARS-CoV-2 infection status: February stress control births, dyads with maternal SARS-CoV-2 infection during pregnancy, and dyads without maternal SARS-CoV-2 infection during pregnancy. Continuous variables were compared using the Kruskal-Wallis test and categorical variables were compared using the Chi-square test. Breastfeeding practice was dichotomized into a binary variable “mostly or exclusively breastfeeding” (combining response categories *all breastmilk* and *mostly breastmilk but some formula*) versus “not mostly or exclusively breastfeeding” (combining response categories *all formula*, *mostly formula but some breastmilk*, and *equal amounts of breastmilk and formula*). Our primary analysis compared this “mostly or exclusively breastfeeding” variable between all SARS-CoV-2 infected vs. uninfected mothers. Responses to additional breastfeeding variables (Table 1) were then compared between SARS-CoV-2 infected and uninfected mothers. Our secondary analysis compared the “mostly or exclusively breastfeeding” variable between SARS-CoV-2 infected and uninfected mothers while separating dyads into two groups: birth during the first wave of the pandemic and birth during the second wave of the pandemic. Our tertiary analysis compared the “mostly or exclusively breastfeeding” variable between SARS-CoV-2 infected vs. uninfected mothers while separating mothers into Latina vs. Not Latina groups.

Logistic regression models estimated the association among maternal SARS-CoV-2 infection status, birth timing during the COVID-19 pandemic (peak vs. post-peak) and mostly or exclusively breastfeeding at 4 months. Models were adjusted for maternal ethnicity only as ethnicity was highly correlated with medical insurance type (*p*<0.0001). All analyses were conducted using R, version 3.6.1 (R Foundation) with a nominal alpha level of 0.05.

## Results

### Demographics

Our study sample included 557 dyads who delivered between February 1, 2020, and April 30, 2021, and enrolled in the COMBO Initiative postnatally. 221 mothers (40%) had a SARS-CoV-2 infection during pregnancy, and the majority (49%) were infected in the 3^rd^ trimester (Table 2). 267 (48%) had no history of SARS-CoV-2 infection, and 69 (12%) were birth-stress controls who delivered in February 2020 (Table 2). Mothers had a median age of 32 years (interquartile range [IQR] 28-36 years) at the time of delivery, and 48% were publicly insured or uninsured. Infants were 46% female with median gestational age at birth of 39.0 weeks (IQR 38.0-39.7 weeks). Compared to uninfected mothers and those who delivered in February 2020, mothers with prenatal SARS-CoV-2 infections were more likely to report Latina ethnicity (73% infected vs. 42% uninfected vs. 56% February stress control, *p*<0.001) and be publicly insured (57% infected vs. 39% uninfected vs. 49% February stress control, *p*<0.001).

**Table 2.**
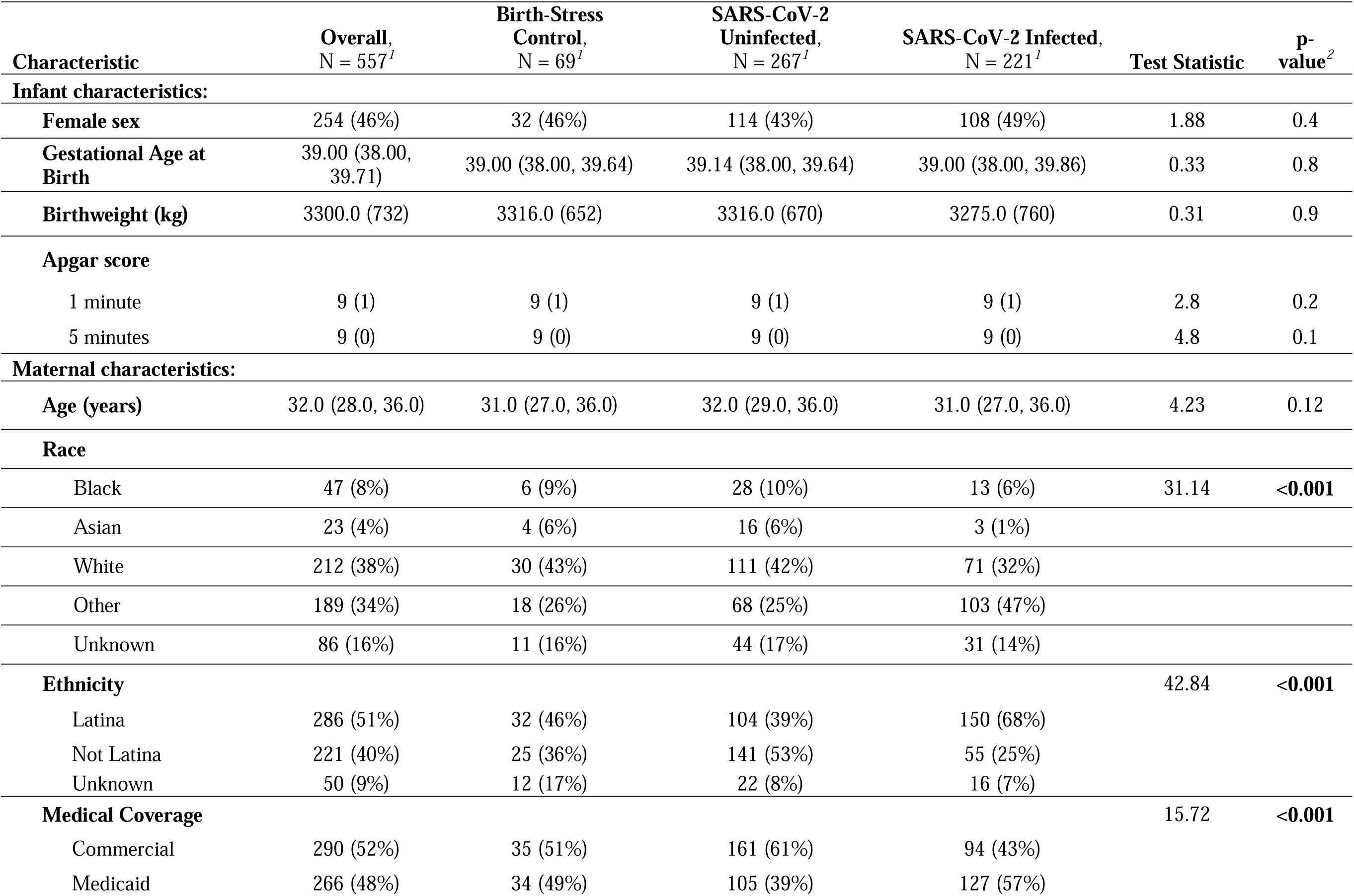
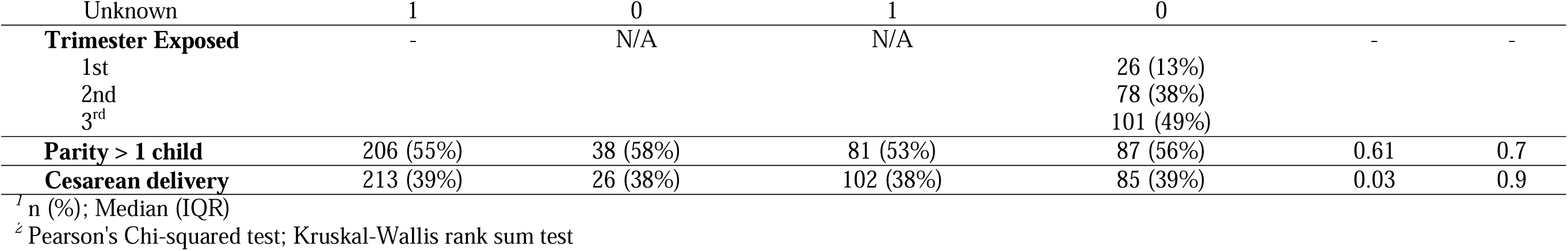
Descriptive statistics of the sample by enrollment group.

### Breastfeeding Practices

Because data from the February stress control group was only obtained at the 4-month time point, one Chi-squared test compared maternal responses to the question “How are you feeding the baby?” among all three enrollment groups at 4 months, and this analysis was repeated at the 1 month, 2 month, 4 month, and 6 month time points after excluding February stress controls. At the 4-month time point, a significantly lower proportion of mothers with history of prenatal SARS-CoV-2 infection reported mostly or exclusively breastfeeding (40%) compared to uninfected mothers (58%) and February stress controls (58%) (*χ^2^*=7.80; *p*=0.020). After excluding February stress controls, a significantly lower proportion of mothers with a history of SARS-CoV-2 infection than mothers with no history of infection reported mostly or exclusively breastfeeding at 1 month (51% infected, 66% uninfected; *χ^2^*=4.05; *p*=0.044), 2 months (46% infected, 62% uninfected; *χ^2^*=7.45; *p*=0.006), and 4 months (41% infected, 56% uninfected; *χ^2^*=5.73; *p*=0.017), as shown in Figure 1.

**Figure 1.**
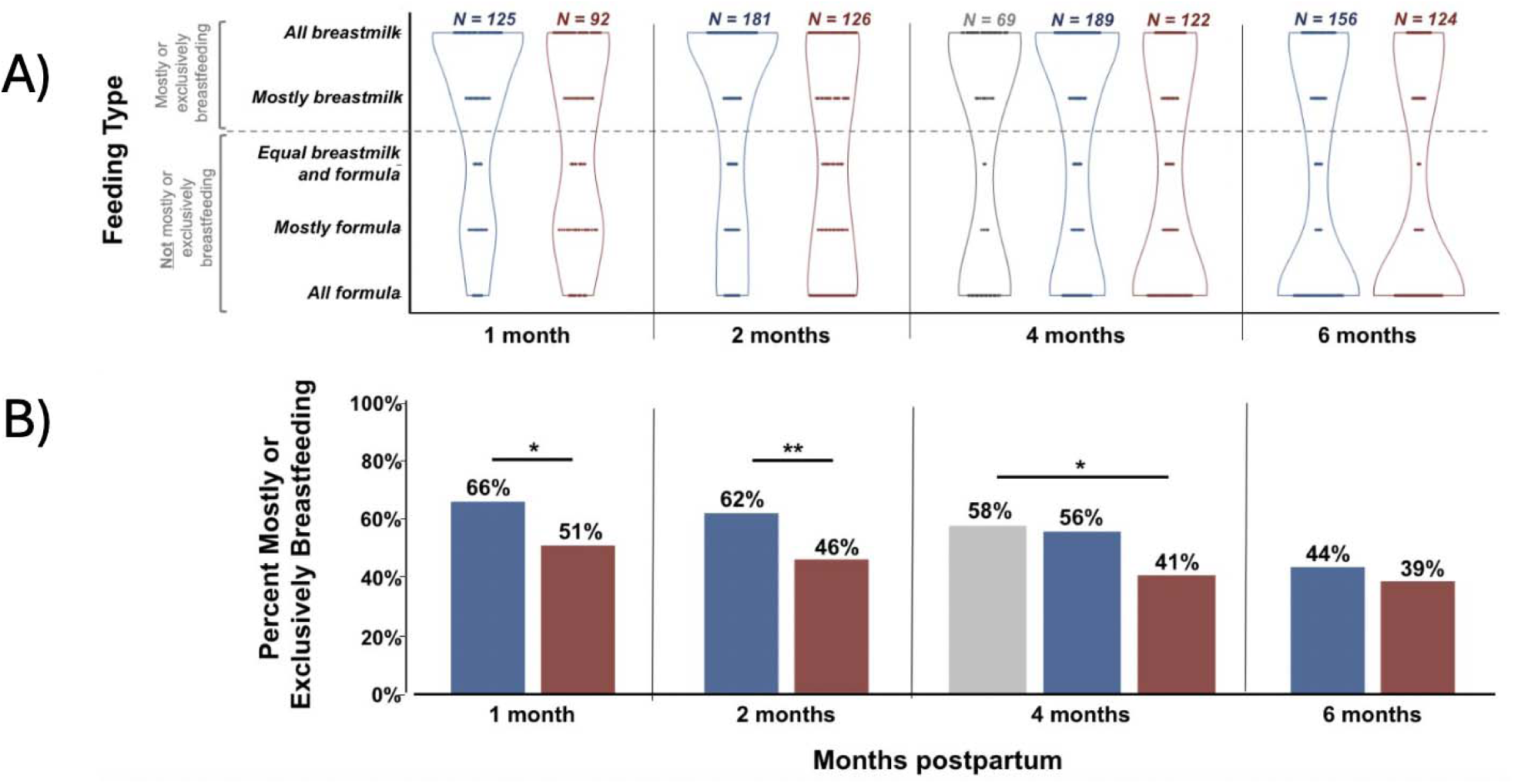
(A) Distribution of feeding type responses by mothers at the 1, 2, 4, and 6 month time points in February Stress Controls (Grey), SARS-CoV-2 uninfected (Blue), and SARS-CoV-2 infected (Red) mothers. Y-axis illustrates how the five feeding type response categories were combined into the binary feeding type variable used in subsequent analyses and shown in part B. (B) Proportion of SARS-CoV-2 uninfected (Blue) and infected (Red) mothers reporting mostly or exclusively breastfeeding their infants at the 1, 2, 4, and 6 month time points. *indicates *p*<0.05, **indicates *p*<0.01.

### Differences in self-reported variables related to breastfeeding

At the time of their first survey, there was no significant difference between SARS-CoV-2 infected vs. uninfected mothers in the proportion who self-reported that they had initially planned to breastfeed, or in their mean ratings of breastfeeding education or support in the hospital, or communication with healthcare providers with respect to breastfeeding. In SARS-CoV-2 infected mothers, the proportion reporting mostly or exclusively breastfeeding was compared between trimester of SARS-CoV-2 infection groups at each time point, and there were no significant differences at any time point (1 month *p*=0.28; 2 months *p*=0.28; 4 months *p*=0.80; 6 months *p*=0.71). The total number of self-reported concerns related to breastfeeding was summed for each participant at each timepoint, and at 1 month the uninfected mothers reported more concerns than infected mothers (median 1.0, IQR 0.0-2.0 infected; median 2.0, IQR 1.0-3.0 uninfected; Wilcox test statistic = 4549.50; *p*=0.007). There was no significant difference by infection group in the median number of concerns at the 2, 4, or 6 month timepoints. A smaller proportion of SARS-CoV-2 infected mothers as compared to uninfected mothers reported being seen by a lactation consultant in the hospital after delivery (58% infected, 76% uninfected; *χ*^2^=13.98; *p*<0.001).

### Breastfeeding practices and birth timing

Figure 2A shows the percentage of SARS-CoV-2 infected vs. uninfected mothers who reported mostly or exclusively breastfeeding at the 1 month, 2 month, 4 month, and 6 month time points, separated by whether infants were born during the first wave (March 2020 through August 2020) or the second wave (September 2020 through April 2021) of the COVID-19 pandemic. Among mothers who delivered during the second wave of the pandemic, a significantly lower proportion of mothers with a history of prenatal SARS-CoV-2 infection reported mostly or exclusively breastfeeding when compared to uninfected mothers at the 1 month (43% infected, 67% uninfected; *χ^2^*=5.23; *p*=0.022) and 2 month (38% infected, 64% uninfected; *χ^2^*=7.78; *p*=0.005) time points. There were no statistically significant differences in breastfeeding rates between SARS-CoV-2 infected vs. uninfected mothers who delivered during the pandemic’s first wave.

**Figure 2.**
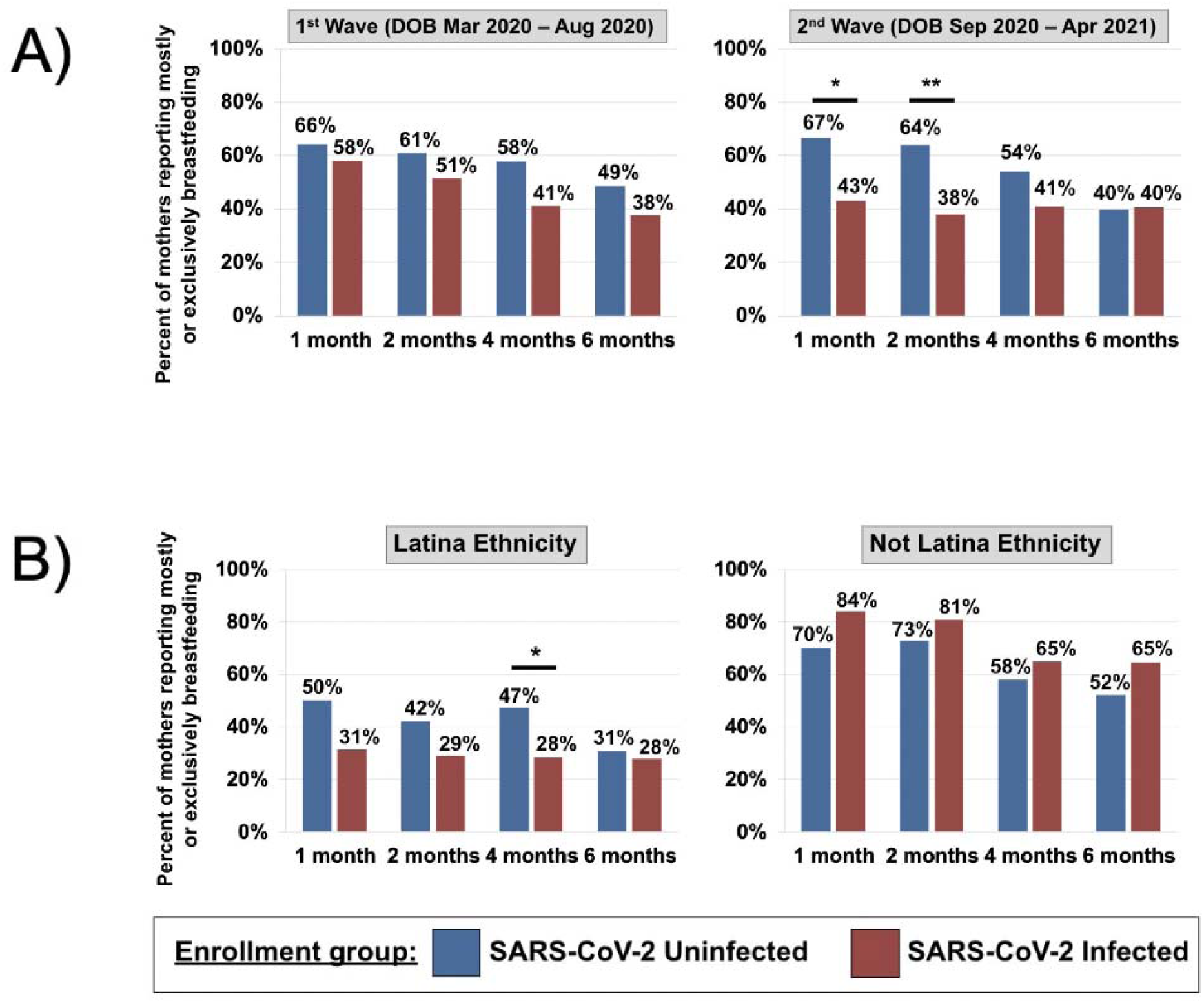
(A) Proportion of SARS-CoV-2 uninfected (Blue) and infected (Red) mothers reporting mostly or exclusively breastfeeding their infants at the 1, 2, 4, and 6 month time points, separated by infant date of birth during the first vs. second wave of the pandemic. *indicates *p*<0.05, **indicates *p*<0.01. (B) Proportion of SARS-CoV-2 uninfected (Blue) and infected (Red) mothers reporting mostly or exclusively breastfeeding their infants at the 1, 2, 4, and 6 month time points, separated by self-reported maternal ethnicity. *indicates p<0.05.

### Breastfeeding practices and maternal ethnicity

Figure 2B shows the percentage of SARS-CoV-2 infected vs. uninfected mothers who reported mostly or exclusively breastfeeding at the 1 month, 2 month, 4 month, and 6 month time points, separated by self-reported maternal ethnicity (Latina vs. Not Latina). In the Latina group, a significantly lower proportion of SARS-CoV-2 infected mothers reported mostly or exclusively breastfeeding when compared to uninfected mothers at the 4 month time point (28% infected, 47% uninfected; *χ^2^*=7.53; *p*=0.023). There were no statistically significant differences in breastfeeding rates at any time point between SARS-CoV-2 infected vs. uninfected mothers in the Not Latina group, and there were no significant differences in the Latina group at the 1 month, 2 month, or 6 month time points.

### Logistic Regression Model

A logistic regression model was used to predict mostly or exclusively breastfeeding at 4-months as this was only time point that included all three cohorts. Predictor variables included maternal prenatal SARS-CoV-2 infection, maternal ethnicity, and birth timing during the pandemic (Table 3). In this model, prenatal SARS-CoV-2 infection was not significantly associated with mostly or exclusively breastfeeding at the 4-month time point when adjusting for maternal ethnicity and birth timing. Birth timing (post-peak pandemic) was negatively associated with mostly or exclusively breastfeeding when controlling for SARS-CoV-2 exposure status and maternal ethnicity (adjusted odds ratio [AOR] 0.43, 95% CI 0.19-0.95, *p*=0.041). Maternal ethnicity (not Latina) was positively associated with mostly or exclusively breastfeeding when controlling for SARS-CoV-2 exposure status and birth timing (AOR 2.83, 95% CI 1.24-6.62, *p*=0.014).

**Table 3.**
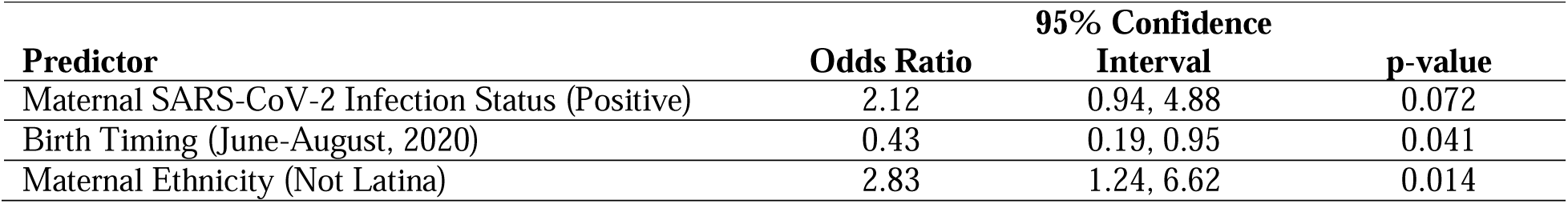
Logistic regression models with maternal SARS-CoV-2 infection status, birth timing, and maternal ethnicity as predictors for mostly or exclusively breastfeeding at 4 months postpartum.

## Discussion

In this observational prospective cohort study, we investigated breastfeeding outcomes after hospital discharge in mother-infant dyads in NYC during the COVID-19 pandemic. Specifically, we assessed differences in breastfeeding outcomes from discharge to 6 months in mothers who were infected with SARS-CoV-2 during pregnancy vs. uninfected, while considering birth timing during the first vs. second wave of the pandemic and maternal ethnicity.

Our findings indicated that mothers exposed to prenatal SARS-CoV-2 infection were more likely to identify as Latina compared to mothers who were unexposed or gave birth in February of 2020. Additionally, mothers who were infected with SARS-CoV-2 during pregnancy were less likely to report mostly or exclusively breastfeeding their infants at 1, 2, and 4 months of infant age compared to uninfected mothers. Results from factors we assessed that may have influenced lower breastfeeding rates in infected mothers are unclear. Interestingly, infected mothers reported fewer concerns related to breastfeeding at the time of their first survey than uninfected mothers but were less likely to report having been seen by a lactation consultant in the hospital after delivery. SARS-CoV-2 infected vs. uninfected mothers did not differ in their initial plans to breastfeed, or their ratings of breastfeeding education, support, and breastfeeding-related communication with healthcare providers while in the hospital after delivery. To assess the possibility that infected mothers were experiencing illness after delivery that may have interfered with their ability to breastfeed, we compared breastfeeding rates across the trimester of SARS-CoV-2 infection and found no difference in rates.

When we bisected participants depending on the wave of the pandemic in NYC during which infants were born, we found that SARS-CoV-2 infected women who delivered during the second wave were less likely to report mostly or exclusively breastfeeding than uninfected women at 1 and 2 months postpartum. No difference in mostly or exclusively breastfeeding rates between infected and uninfected women was found in the group of participants who delivered during the first wave.

Collectively, fewer mothers who had SARS-CoV-2 infection during pregnancy and self-identified as Latina reported mostly or exclusively breastfeeding infants at 4 months of age as compared to non-Latina mothers. This difference reflects previously studied ethnic disparities in health and breastfeeding outcomes^13–16^ and suggests that exposure to SARS-CoV-2 infection had a “double-hit” negative impact on breastfeeding outcomes in combination with documented disparities.^17^

We discuss themes that emerged from our findings with implications for current maternal and child health efforts, including cultural differences in breastfeeding practices, the effect of social determinants on health outcomes, and the disadvantages of decreased support systems during public health emergencies such as the COVID-19 restrictions.

### Las Dos Cosas and cultural differences in breastfeeding practice

Our hospital system serves a predominantly Dominican community in Northern Manhattan.^12^ Prior studies have reported that Latina mothers initiate breastfeeding at high rates but subsequently have lower rates by 6 and 12 months compared to other ethnic groups in the U.S.^13^ Latina mothers are more likely than other ethnic groups to supplement with formula as early as the second day of life.^13^ This phenomenon is described in the literature as “las dos cosas”^14–16^ and likely has a negative impact on breastfeeding to 6 months of age by compromising breastmilk production.^16^ A previous study from our hospital system^12^ found high rates of formula supplementation in the first 4-6 weeks of life. Hypotheses for this early introduction of formula include a desire for the infant to receive both the benefits of breastmilk and “vitamins” in formula, insufficient milk supply/breastfeeding difficulties, longer infant satiety, family/cultural beliefs about infant weight, and plans to return to work.^14,15^ Cultural differences in breastfeeding practice likely contributed to the decrease in mostly or exclusively breastfeeding at 4 months of age in Latina mothers infected with SARS-CoV-2 during pregnancy.

### Social Determinants of Health

The COVID-19 pandemic disproportionately affected historically marginalized groups, including Latina communities, due to the structural, societal and economic causes of health disparities.^18,19^ Social distancing and working from home were not feasible for many patients who lived in housing conditions or worked in occupations that put them at higher risk for contracting COVID-19.^18^ Such structural inequalities were exacerbated throughout the pandemic, and likely contributed to more Latina mothers being in the infected group. Similar factors could explain differences in breastfeeding rates between infected and uninfected mothers which were only significant in those who gave birth during the second wave of the pandemic in NYC, after stay-at-home orders were lifted. Given demographic differences, more mothers in the infected group than in the uninfected group may have had to return to work during the second wave.

### Decreased professional and social support

In addition to existing health disparities, the disruption of traditional provider and social support systems during the pandemic may have contributed to decreased breastfeeding rates in SARS-CoV-2 infected mothers. Prior studies showed that breastfeeding outcomes are most effective when in-person support is provided by professionals, peers, or a combination.^20^ A CDC survey of hospitals during the pandemic found that most shortened the birth hospitalization to less than 48 hours, and some decreased lactation services.^9^ Shorter lengths of stay and decreased availability of in-person lactation consultants may have translated into decreased time focused on breastfeeding support for postpartum mothers with SARS-CoV-2 infection, consistent with our finding that infected mothers were less likely to report having been seen by a lactation consultant in the hospital.

Likewise, decreased social support and increased isolation during the pandemic also may have impacted breastfeeding. Prior to the pandemic, our hospital system allowed for eight visitors in a single postpartum room and four visitors in a shared postpartum room. At the onset of the pandemic, there were visitor policy changes for the birth hospitalization. Beginning on March 17, 2020, one visitor was allowed per laboring mother on the Postpartum Unit. On March 22, 2020, the policy changed to prohibit all visitors from the Postpartum Unit. Restrictions were eased to allow one support person and one trained doula on April 30, 2020. Outside of the hospital, in-person breastfeeding and new parent support groups were cancelled. Such cancellations combined with decreased support from friends and family may have contributed to earlier cessation of breastfeeding.^9^ A survey of pregnant and breastfeeding women in Belgium during the first peak of the pandemic in April 2020 found that changes in medical counseling and lack of social support negatively affected women who had not breastfed before and those in the early postpartum period during that time.^21^ Another study of breastfeeding mothers in the US in March to June 2020 found that women reported less familial and peer-to-peer support available at that time and which impaired their breastfeeding experience.^20^

### Limitations

As this was a cross sectional study, our results can only infer correlation and not causation. Additionally, results from this single hospital system may not be generalizable to other demographics. Our surveys were unable to fully elucidate factors during the pandemic period which may have resulted in decreased breastfeeding, particularly while in the hospital. For example, while official hospital policy was to continue rooming mothers and babies together and encourage direct breastfeeding^11^, guidance about maternal/newborn isolation and breastfeeding from staff may have differed given uncertainty early in the pandemic. Additionally, our surveys did not elucidate the reasons for difficulty with or stopping breastfeeding and further research in this area would be beneficial.

## Conclusions for Practice

Our study found that maternal self-reported breastfeeding practices were affected by the COVID-19 pandemic. Mothers with prenatal SARS-CoV-2 infection were less likely to breastfeed at 1, 2, and 4 months of infant age than mothers without prenatal SARS-CoV-2 infection. This difference was particularly evident in mothers who delivered during the second wave and was present in our sample despite our hospital system’s encouragement of newborns rooming-in and direct breastfeeding. Mothers of Latina ethnicity who had a SARS-CoV-2 infection during pregnancy were especially likely to report less breastfeeding. Although the official pandemic has ended, these results highlight the need to protect and increase breastfeeding support during potential future disruptive public health emergencies and to increase accessibility of these resources to those in groups historically underserved in healthcare.

## Data Availability

All data produced in the present study are available upon reasonable request to the authors.

## Acknowledgments

We thank the entire COMBO study team and all the COMBO families for their help with this project.

